# Lockdown measures in response to COVID-19 in Sub-Saharan Africa: A rapid study of nine countries

**DOI:** 10.1101/2020.07.09.20149054

**Authors:** Najmul Haider, Abdinasir Yusuf Osman, Audrey Gadzekpo, George O. Akpede, Danny Asogun, Rashid Ansumana, Richard Lessells, Palwasha Khan, Muzamil Mahdi Abdel Hamid, Dorothy Yeboah-Manu, Leonard Mboera, Elizabeth H Shayo, Blandina T Mbaga, Mark Urassa, David Musoke, Nathan Kapata, Rashida Abbas Ferrand, Pascalina Chanda-Kapata, Florian Stigler, Thomas Czypionka, Richard Kock, David McCoy

## Abstract

Lockdown measures have been introduced worldwide to contain the transmission of COVID-19. This paper defines the term lockdown and describes the design, timing and implementation of lockdown in nine countries in Sub Saharan Africa: Ghana, Nigeria, South Africa, Sierra Leone, Sudan, Tanzania, Uganda, Zambia and Zimbabwe. It also discusses the manner in which lockdown is enforced, the need to mitigate the harms of lockdown, and the association between lockdown and the reported number of COVID-19 cases and deaths. While there are some commonalities in the implementation of lockdown, a more notable finding is the variation in the design, timing and implementation of lockdown measures across the nine countries. We found that the number of reported cases is heavily dependent on the number of tests done, and that testing rates ranged from 9 to 21,261 per million population. The reported number of COVID-19 deaths per million population also varies, but is generally low when compared to countries in Europe and North America. While lockdown measures may have helped inhibit some community transmission, the pattern and nature of the epidemic remains unclear. Of concern are signs of lockdown harming health by affecting the functioning of the health system and causing social and economic harms. This paper highlights the need for inter-sectoral and trans-disciplinary research capable of providing a rigorous and holistic assessment of the harms and benefits of lockdown.

## Introduction

Thus far, the SARS-CoV-2 (COVID-19) pandemic which was officially declared by the World Health Organization (WHO) in March 2020 appears to have mainly affected wealthier countries. As of June 2 2020, 65% of all reported cases globally were from countries in Europe and North America [1]. Although it was predicted that Africa’s epidemic would be delayed compared to Europe and North America due to the relatively lower risk of cases being imported from China,[2,3] the number and proportion of reported cases in Africa remains low, amounting to only 157,254 cases or 2.5% of the global total at that time (with South Africa, Egypt and Nigeria recording the most cases)[1].

The reported data from Africa are likely to underestimate the true magnitude of the pandemic due to under-detection of cases, as well as under-reporting of detected cases. However, the experience thus far suggests that the disease is less severe in Africa compared to Europe, North America, Asia and South America[4]. On May 22^nd^, the World Health Organisation even stated that the pandemic “appears to be taking a different pathway in Africa” and that “so far, Africa has not experienced the high mortality seen in some parts of the world” [5]. Postulated reasons for this include Africa’s younger demographic, higher average temperatures and the existence of higher levels of pre-existing immunity [6].

The high transmissibility of COVID-19 and the fact that asymptomatic or pre-symptomatic individuals may be contagious [7] has meant that standard communicable disease (CD) control measures involving active case detection, contact tracing and selective isolation and quarantine may be insufficient to bring transmission under control, especially when infection rates are comparatively high. As a consequence, many countries have deployed community-wide ‘lockdown’ measures to reverse exponential epidemic growth trajectories.

Although the term ‘lockdown’ is now ubiquitous, it is not well-defined. There are also no clear definitions of commonly used adjectives for the term such as ‘total lockdown’ and ‘partial lockdown’; and ‘hard’ or a ‘soft’ lockdown. Indeed, the WHO’s reference to ‘*so-called* lockdown measures’ indicates the absence of a clear and universally-accepted definition of the term lockdown[8]. Given its widespread use and importance, we have come up with a definition of ‘lockdown’ using a two-by-two matrix based on whether measures are compulsory or voluntary; and whether they are targeted at individuals or applied to a general population (Table 1).

**Table 1:** Categorisation of communicable disease control measures

|  | **Voluntary / Advisory** | **Compulsory / Mandatory** |
| --- | --- | --- |
| **Targeted or focused around individuals** | Encourage actions and behaviours amongst identified or suspected cases, and contacts of cases<br>- Hygiene measures (including wearing facemasks)<br>- Physical distancing<br>- Isolation or quarantine | Actions and behaviours required of identified or suspected cases, and contacts of cases<br>- Hygiene measures (including wearing facemasks)<br>- Physical distancing (including working from home)<br>- Isolation or quarantine |
| **Measures applied to a general population** | Encourage actions and populations in general population(s)<br>- Hygiene measures (including wearing facemasks)<br>- Physical distancing<br>- Refrain from non-essential travel<br>- Avoid gatherings of people | Actions and behaviours required of general populations<br>- Hygiene measures (including wearing facemasks)<br>- Physical distancing (including working from home)<br>**Geographic containment**<br>**Home confinement**<br>**Prohibition of gatherings and closure of certain establishments and premises** |

Using this matrix, we define lockdown as *a set of measures aimed at reducing transmission of COVID-19 that are mandatory, applied indiscriminately to a general population and involve some restrictions on normal social and economic life*. This definition has been refined from an earlier published version[9] and excludes measures that are compulsory but targeted at individuals or high-risk population groups; and population-wide measures which are compulsory but do not involve any significant restriction on freedom and normal social and economic life such as being required to war facemasks (FMs) in public or having to abide by physical distancing (PD) stipulations in public.

The boundaries between some of the four quadrants may be blurred and open to some varied interpretation, but based on this definition we have identified three interventions in the bottom right quadrant of the matrix which constitute ‘lockdown’: (i) geographic containment; (ii) home confinement; and (iii) prohibition of gatherings and closure of certain establishments and premises.

Geographic containment is a type of lockdown measure that is perhaps best associated with the decision of Chinese authorities to stop the movement of people in and out of Wuhan city. It is designed to prevent epidemic hotspots from contaminating other parts of a country or region. Exemptions will usually be made to ensure the flow of food and other essential commodities in and out of a locked down area, and there may be minimal or absent restrictions for people travelling *into* an area that has been put into lockdown. A *cordon sanitaire* may accompany geographic containment. This term refers to the creation of a buffer zone around an area experiencing an epidemic across which there is movement control and which therefore helps to act as a barrier to disease transmission.

Home confinement refers to requirements placed on a general population to stay at home for prescribed amounts of time. The term ‘curfew’ is sometimes used interchangeably with home confinement, and exemptions are typically made for people whose jobs are considered essential, or for certain permitted activities such as shopping for food or taking exercise.

The third type of lockdown measure is the prohibition of gatherings and the closure of certain social, recreational, educational, religious and economic establishments and premises. This includes the closure of schools, universities, restaurants, cinemas, theatres, churches, mosques, and retail outlets; and the prohibition ore restriction of gatherings of people. As with other lockdown measures, exemptions are common and may include essential businesses and industries staying open; certain types of gatherings being allowed to continue (eg. funerals); or certain premises being kept open for defined groups of people (eg. schools being kept open for the children of essential workers).

A clear and bounded definition of lockdown is important from a research perspective because of the need to monitor its effectiveness and impact on disease control. Furthermore, lockdown poses several threats to health and well-being and may even cause more overall harm than good. At the level of individuals, lockdown can result in psychological and emotional distress; loss of employment and household income; and deprive children of the benefits of schooling (Brooks et al., 2020). These harms are aggravated by the effects of lockdown at the societal level including economic contraction and recession, disruption of supply systems, aggravation of social tensions, and the potential for lockdown to lead to the long-term erosion of human rights and civil liberties.

It is therefore important that public health systems monitor and evaluate the impact of lockdown in terms of epidemic control, and their wider social, economic, health and political impacts. Attention should also be paid to evaluating the existence of measures designed to mitigate the harms of lockdown, such as providing financial and welfare support to vulnerable households and businesses, organising online schooling for children and introducing fiscal measures to keep the economy afloat.

This paper describes the design, timing and implementation of the three types of lockdown in a set of nine countries in SSA: Ghana, Nigeria, South Africa, Sierra Leone, Sudan, Tanzania, Uganda, Zambia and Zimbabwe (see **Table 2 and Fig 1**). It also describes the manner in which lockdown was enforced and the efforts made to mitigate the harms of lockdown.

**Table 2:**
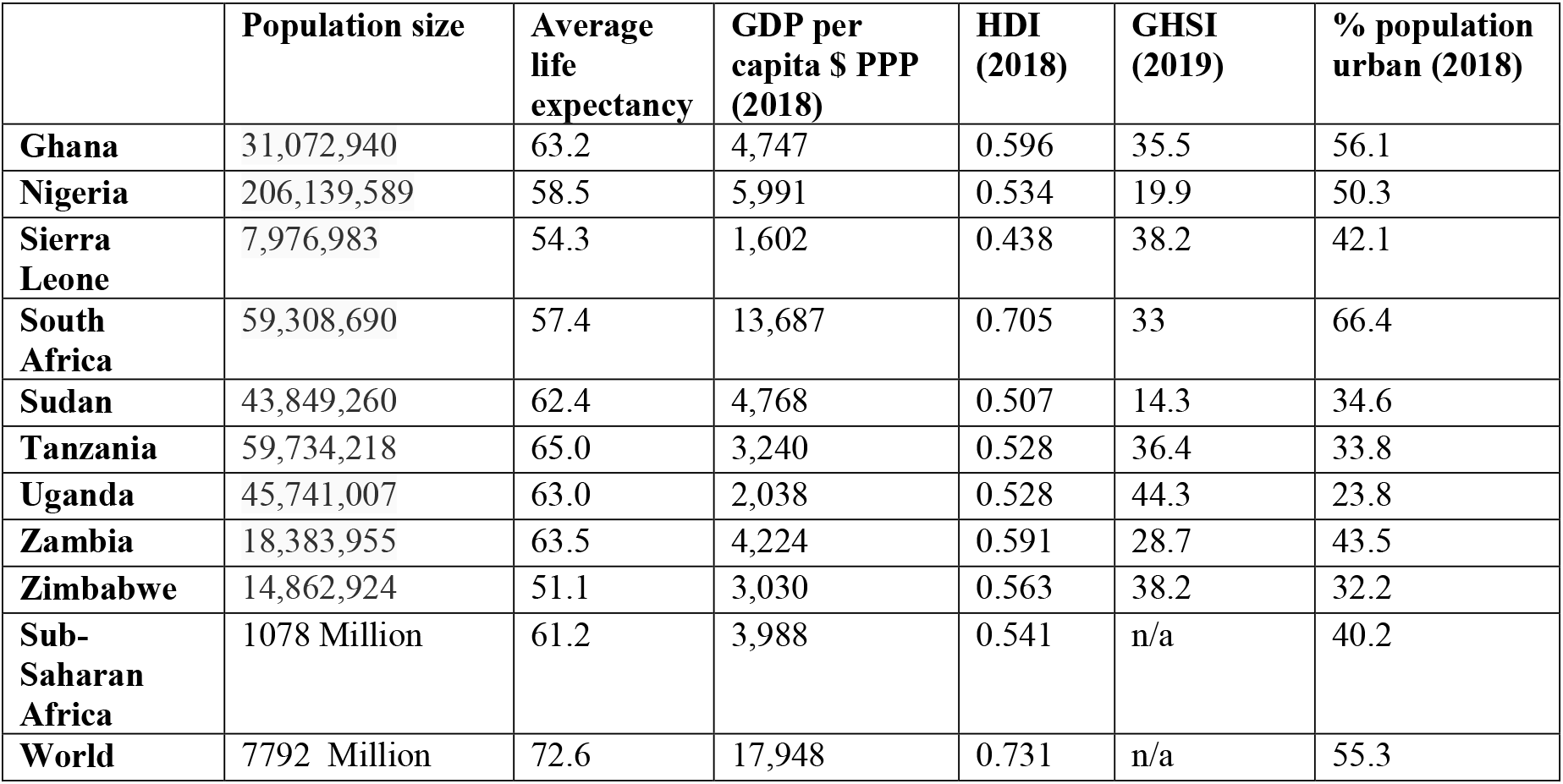
Demographic information of nine sub-Saharan countries

| | Population size | Average life expectancy | GDP per capita \$ PPP (2018) | HDI (2018) | GHSI (2019) | % population urban (2018) |
| --- | --- | --- | --- | --- | --- | --- |
| **Ghana** | 31,072,940 | 63.2 | 4,747 | 0.596 | 35.5 | 56.1 |
| **Nigeria** | 206,139,589 | 58.5 | 5,991 | 0.534 | 19.9 | 50.3 |
| **Sierra Leone** | 7,976,983 | 54.3 | 1,602 | 0.438 | 38.2 | 42.1 |
| **South Africa** | 59,308,690 | 57.4 | 13,687 | 0.705 | 33 | 66.4 |
| **Sudan** | 43,849,260 | 62.4 | 4,768 | 0.507 | 14.3 | 34.6 |
| **Tanzania** | 59,734,218 | 65.0 | 3,240 | 0.528 | 36.4 | 33.8 |
| **Uganda** | 45,741,007 | 63.0 | 2,038 | 0.528 | 44.3 | 23.8 |
| **Zambia** | 18,383,955 | 63.5 | 4,224 | 0.591 | 28.7 | 43.5 |
| **Zimbabwe** | 14,862,924 | 51.1 | 3,030 | 0.563 | 38.2 | 32.2 |
| **Sub-Saharan Africa** | 1078 Million | 61.2 | 3,988 | 0.541 | n/a | 40.2 |
| **World** | 7792 Million | 72.6 | 17,948 | 0.731 | n/a | 55.3 |

**Fig-1:**
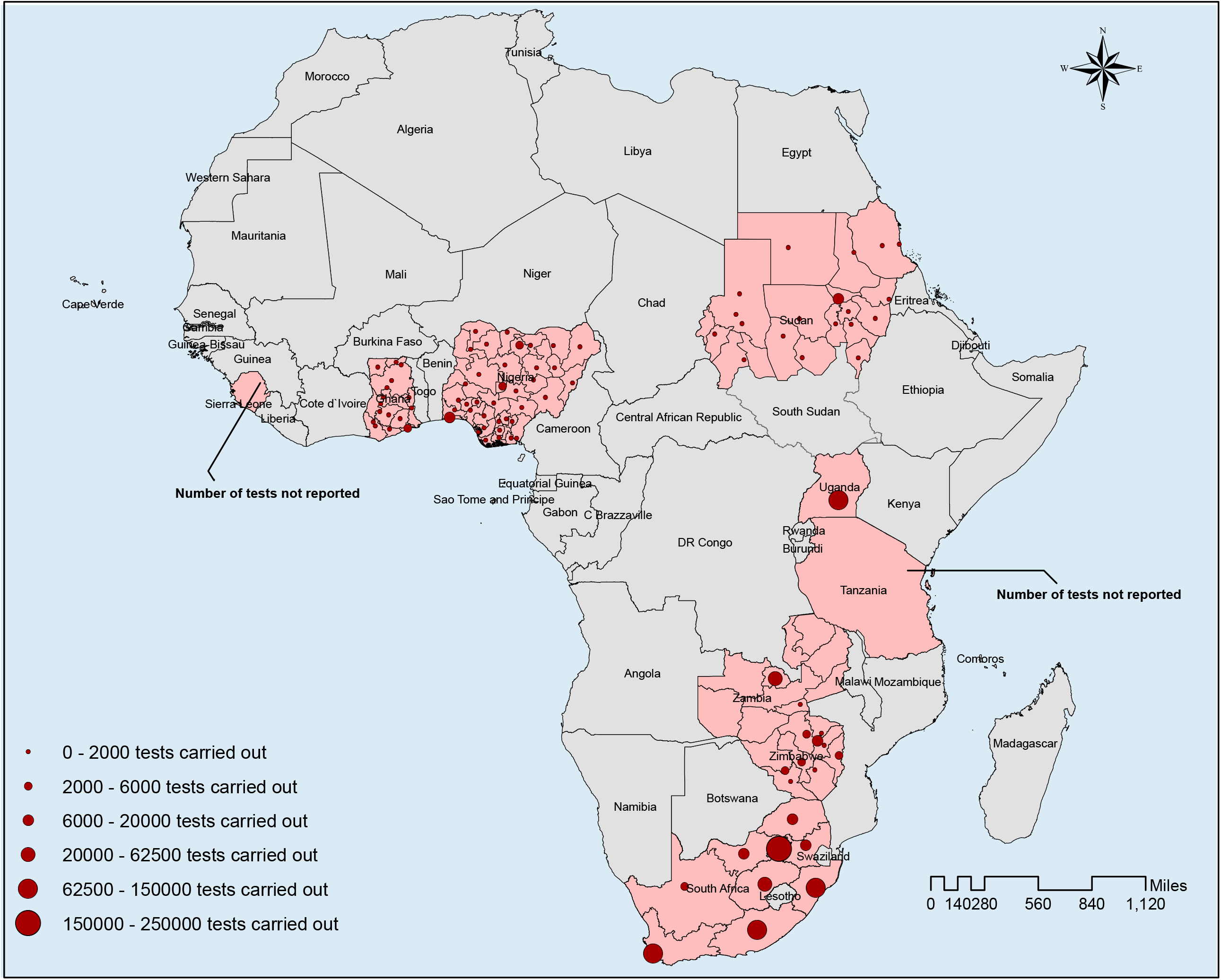
The map with nine countries of Sub-Saharan Africa including the number of samples tested in 7 countries (location of testing canters and number is shown for Nigeria, Ghana, South Africa, Sudan and Zimbabwe and aggregated national number is shown for Uganda and Zambia).

## Methods

The research was conducted by a group of country-based researchers who volunteered to participate in what was designed to be a rapid research exercise. Data were collected by country-based researchers using a semi-structured questionnaire, supplemented with additional data obtained from the WHO and African CDC websites, and other reliable sources on the worldwide web. Due to constraints in space, we provide a summarised description of lockdown and its impacts here. However, the full set of data we collected is available (on request) for other researchers to assess and use for other research studies.

## Results

### 3.1 Overview of lockdown measures

Although there are some commonalities across the nine countries, a more notable finding is the variation in the design, timing and implementation of lockdown measures (see **Table 1** for a tabulated summary). Tanzania lies at one end of the spectrum with minimal use of lockdown measures while South Africa and Uganda were more extensive and strict in their deployment of lockdown measures. In Nigeria’s federal system, devolution of authority to state governments resulted in lockdown being applied differently across different states making it hard to provide an overall description for the country.

One of the commonalities across the nine countries was the suspension of international passenger flights relatively early in the epidemic. Land and sea border crossings were also restricted, with exceptions made for the continued passage of goods and commodities. The geographic containment of areas within countries was less common with only Nigeria and Ghana targeting lockdown measures to selected areas within the country. Most countries have continued with restrictions on international travel, but in Tanzania, the government has started to lift restrictions and removed mandatory quarantine for international arrivals in May (albeit with enhanced screening of passengers upon arrival).

In all of the countries studied, with the exception of Sierra Leone and Tanzania, non-essential businesses, restaurants, cafes and recreational establishments have been closed, albeit with shops and markets being allowed to continue selling food and other essential commodities. In Sierra Leone, all shops and businesses were allowed to stay open except during two 3-day periods of lockdown; while in Tanzania, there has been no closure at all of shops, government offices, parliamentary sessions, religious congregations, restaurants and cafes. All countries, including Sierra Leone and Tanzania, have closed down schools, colleges and universities, and prohibited or restricted public and private gatherings to greater or lesser degrees. For example, in Sierra Leone only gatherings of more than 100 were prohibited; while in Ghana and Tanzania gatherings of more than 25 and 10 people respectively were prohibited.

There is evidence of all countries allowing businesses to reopen from the middle of April and onwards. In Ghana, businesses and other workplaces were allowed to reopen with PD and hygiene protocols in place. In Zambia, although restaurants, gyms and casinos were allowed to re-open on May 8^th^, bars and taverns remained closed. In Zimbabwe, formal sector businesses were allowed to open from 8am to 3pm, but not informal traders and markets. Generally speaking, the prohibition of large gatherings and closure of educational facilities continued to remain in place throughout the month of May 2020.

When it comes to home confinement and the imposition of curfew, South Africa and Uganda implemented the strictest measures. In South Africa a countrywide curfew began at the end of March 2020 and included the prohibition of any exercise outside the home (only essential activities were allowed between 8am and 5pm). Restrictions were loosened slightly in early May with strict home confinement only required between 8pm and 5am and persons being allowed to exercise between 600 and 900 am provided this is done within 5km of home and not in groups. In Uganda, home confinement was imposed from March 30 and was still in place as per June 15, with a strict curfew imposed from 7pm to 6.30am. In Nigeria, a stay at home order in Lagos State, the Federal Capital Territory (Abuja) and Ogun State for about a month was issued by the Federal government, alongside a nationwide curfew from 8pm to 6am. In Ghana, home confinement was limited to the two major metropolitan areas of Accra and Kumasi for a 3-week period. Sudan’s countrywide curfew initially occurred between 6pm and 6am with extended curfew hours from 1 pm to 7 am for the states of Khartoum, Gezira and Gadaref. These extended hours were then applied to the whole country in mid-April. In Sierra Leone a nationwide curfew was implemented from 9pm to 6am, supplemented by two 3-day periods (April 5-7; May 3-5) of 24-hour home confinement. In contrast, home confinement was not implemented in Zambia and Tanzania. Other CD control measures have been implemented in all nine countries. For example, health information campaigns about COVID-19, hygiene measures such as frequent hand washing, use of FMs and ensuring greater physical distancing have been implemented in all countries. Thus in Tanzania, although all shops have remained open, preventive measures such as hand washing with soap, maintaining PD and wearing of FMs have been widely observed.

Standard CD control measures such as active case detection and isolation, coupled with contact tracing and quarantine have been activated, strengthened and carried out to varying degrees of effectiveness in all countries. This included Ghana, Nigeria, Tanzania, Uganda and Zambia screening and quarantining people who had flown into the country prior to the ban on international travel. Diagnostic testing capacity has been expanded. For example, Ghana started with two diagnostic centres based in major research facilities, but has now been expanded its capacity to about 10 diagnostic centres. The use of FMs in public has been compulsory for the whole population in Ghana, Sierra Leone, Uganda, Zambia and Zimbabwe; but only in certain parts of the country in Tanzania and Zimbabwe. In South Africa, FMs are compulsory on public transport. A particular feature of Ghana’s response has been the use of public disinfection campaigns involving vector-control and waste management companies.

### 3.2 Mitigation measures

The degree of harm that may be caused by lockdown is mainly influenced by the breadth, depth and length of the different lockdown measures. Other factors may also shape the experience of lockdown including the structure of the economy, pre-existing levels of poverty and financial insecurity, the capacity of pre-existing social welfare services, levels of fear about the virus, public trust in government, the degree of social solidarity in society (which influences how people experience lockdown measures psychologically and emotionally), and the deployment of specific mitigation measures.

Most countries have implemented a variety of measures to support poor households as well as small, micro and medium enterprises. In Ghana, a GH¢1.2 billion (about US$200m) Coronavirus Alleviation Programme established by the government to support affected households and businesses, was complemented by donations from private sector companies, non-governmental organisations and civil societies. In South Africa, a social relief and economic support package of R500 billion (about US$3 billion) was established to provide additional welfare and emergency water, sanitation, and shelter services, among other things. In Uganda, the government has distributed food to vulnerable households (but only in Kampala and neighboring towns in the central region). Government support for poorer households has been more limited or absent in Sierra Leone, Tanzania and Zambia, where lockdown measures have been less extensive. In Zimbabwe, cash transfers have been used to support vulnerable groups. [10]

Indirect forms of support to vulnerable households has also been provided. For example, in South Africa, landlords were prohibited from evicting any person from their place of residence (whether formal or informal) for the duration of the lockdown. In Tanzania, water and electricity agencies were ordered not to make any disconnections during the lockdown; while in Ghana, the government gave all citizens three months free water; and electricity was provided free for low-income consumers and at half cost for all other consumers in April, May and June 2020. In Nigeria, the National Electricity Power Agency increased the number of hours of electricity available per day to consumers and the price of fuel/petrol was reduced.

Some countries have also implemented interventions to support the economy. South Africa’s larger economy and greater fiscal flexibility allowed it implement a social relief and economic support package that was worth about 10% of its GDP. In Ghana, the International Monetary Fund (IMF) and private banking sector have intervened to keep credit flowing. In Uganda, the Central bank offered support to commercial banks to continue operating, while a loan of US$491.5m from the IMF’s Rapid Credit Facility was approved to support the economy.

### 3.3 Lockdown and the epidemic

The relationship between the mix of lockdown measures and the number of reported COVID-19 cases and deaths for each of the nine countries are shown in **Fig 2**. The graphs show no obvious pattern. The number of reported cases is heavily dependent on the number of tests done, and the available data shows a considerable variation in testing rates ranging from 9 tests per million population in Sudan to 21,261 tests per million population in South Africa (**Fig 1**).

**Fig-2:**
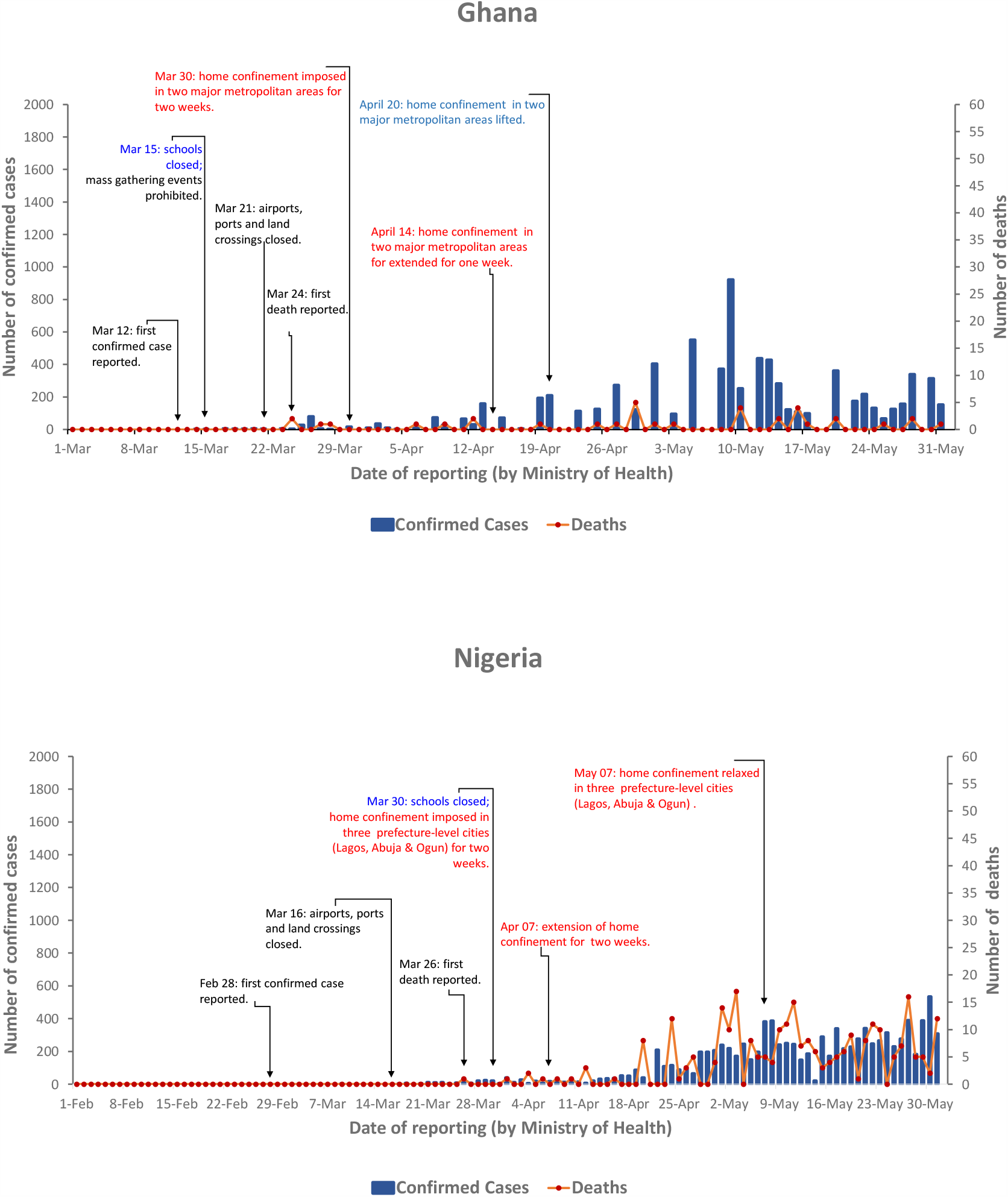
Different measures of Lockdown taken in 9 Sub-Saharan African countries. The daily reported cases are presented in left hand y-axis and death cases are at right hand y-axis. Please notice that axis has different values.

## Discussion

Drawing conclusions about the impact of lockdown measures on COVID-19 transmission is difficult for several reasons. Firstly, the true pattern of the epidemic cannot be readily deduced from the number and pattern of reported cases because many infections are undetected and some may be unreported. Secondly, it is difficult to accurately assess the extent to which the lockdown measures have been properly implemented and adhered to in different parts of the country and over time. Finally, it is not possible to disentangle the effects of different lockdown measures from each other as well as from other CD control measures.

Nonetheless, logic and evidence from elsewhere indicates that lockdown measures will have helped lower the effective reproduction rate of the virus in these nine countries. In particular, geographic containment, travel restrictions and the prohibition of large gatherings should have inhibited community transmission. The relatively large percentage of people living in rural areas where there is naturally more PD and less population mixing may also have contributed to preventing the large epidemic spikes witnessed in other countries.

At the same time, it would be reasonable to think that lockdown measures could help increase COVID-19 transmission in the large and dense informal or semi-formal settlements of Africa. In Accra, for example, about 53% of Accra’s households live in a single room, while an even larger proportion relies on public toilets. where PD and home confinement cannot be practically observed. There are also reports from Zimbabwe that over-crowded and poorly managed quarantine centers have acted as hotspots of transmission.

Although COVID-19 appears to be causing some severe illness in the SSA context, it does not appear to be as severe as it has been in some countries. Meanwhile, concern about the ‘collateral damage’ of lockdown measures is increasing. The World Bank predicted in April 2020 that the economy across the continent could contract by up as much as 5.1% due to a decline in output growth among the region’s key trading partners, a fall in commodity prices, reduced tourism activity, as well as lower levels of foreign direct investment and remittances. Countries dependent on oil exports and mining were expected to be hit hardest[11]. The collapse of the international tourist industry will be felt hard in Sudan, Uganda and Tanzania where tourism makes up a high proportion of overall national income.

Of the nine countries studied here, South Africa stands apart with its relatively large economy (see Table 4) allowing the government greater fiscal space with which to support vulnerable households and the economy. Most of the other countries are much more vulnerable to the negative economic effects of lockdown, and the role of the IMF in enabling countries to access credit and default on debt payments will be crucial in the coming months. Poor households will be hit hardest, especially in urban and periurban areas where many are dependent on daily wage labour in the informal economy and where it is not possible to live off a subsistence economy. Furthermore, the number of poor households will grow as the formal economy contracts and unemployment levels rise. Although rural areas have been less affected by both the virus and lockdown measures, rural households will be affected by a reduction in remittances from family members working in urban areas.

**Table 3:**
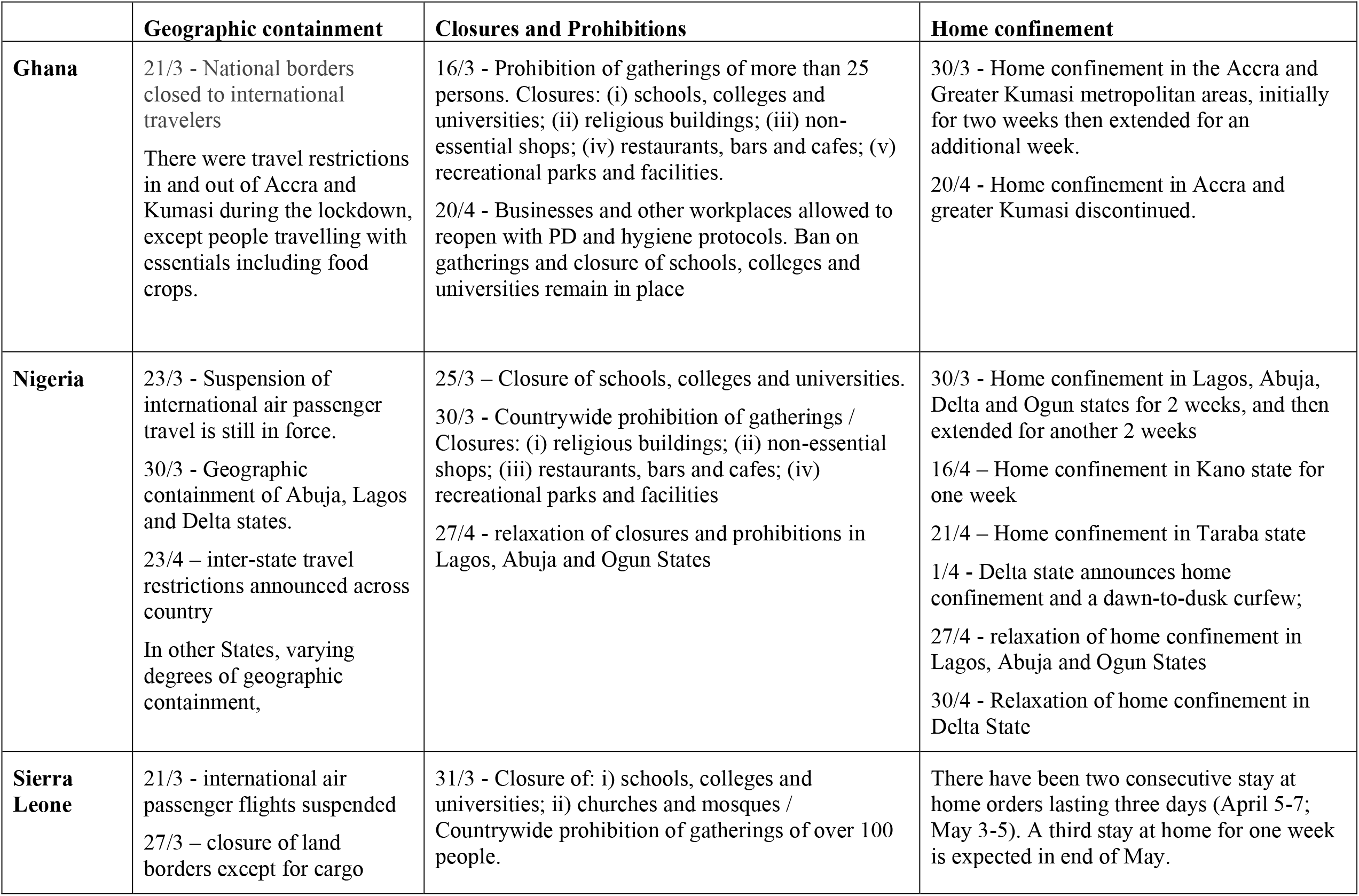
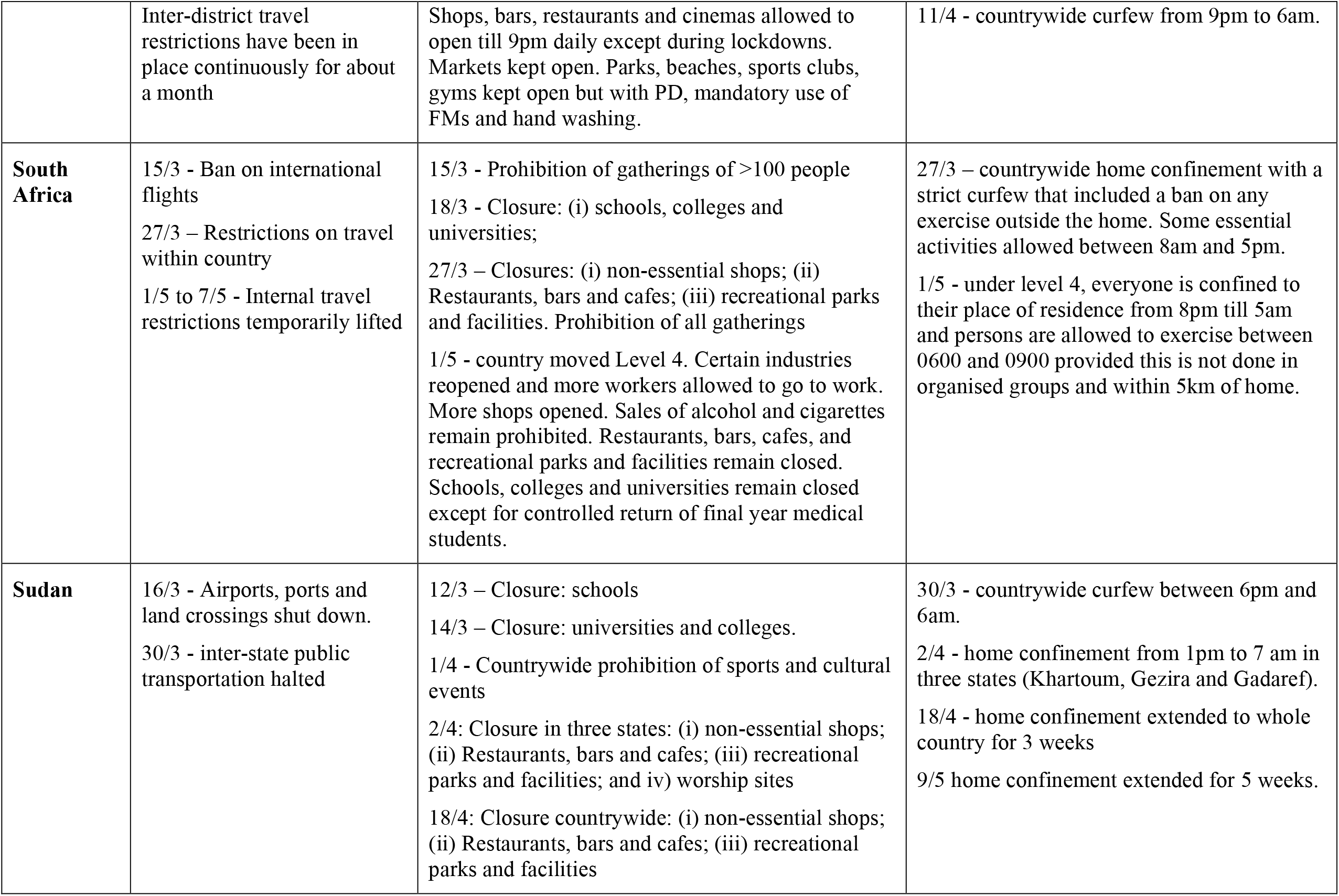
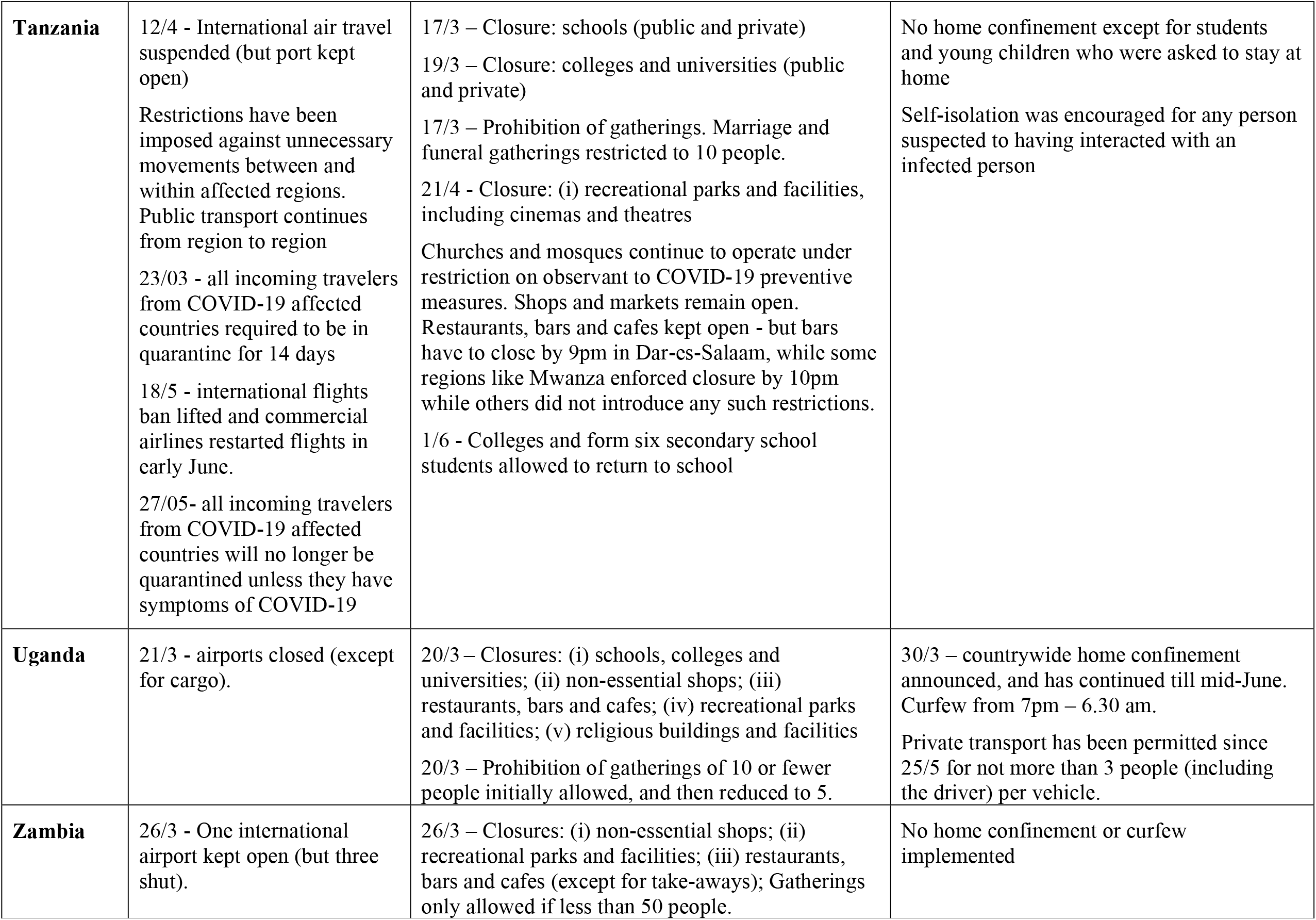
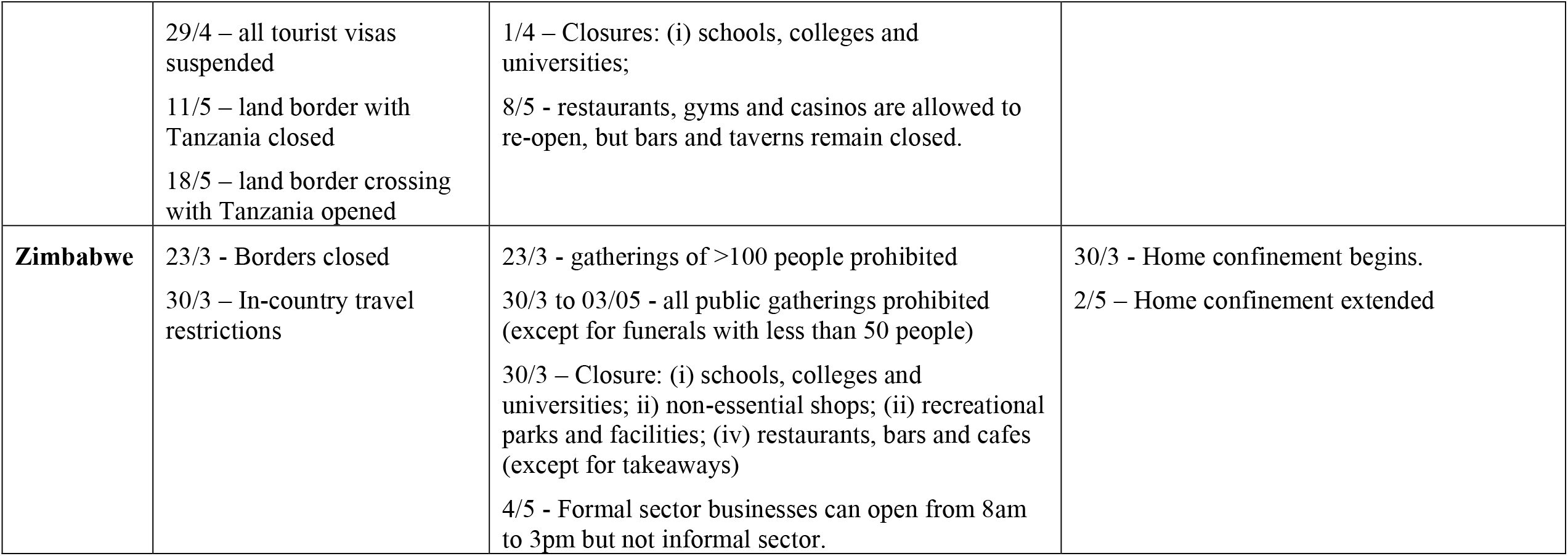
Summary of lockdown measures

**Table 4:** Reported cases, tests and deaths (as of June 20, 2020)[1]

|  | **Total reported COVID-19 cases per million population** | **Total reported COVID-19 tests per million population** | **Total reported COVID-19 deaths per million population** |
| --- | --- | --- | --- |
| **Ghana** | 425 | 8,417 | 2 |
| **Nigeria** | 93 | 527 | 2 |
| **Sierra Leone** | 164 | NA | 7 |
| **South Africa** | 1,480 | 21,261 | 31 |
| **Sudan** | 190 | 9 | 12 |
| **Tanzania** | 9 | NA | 0.4 |
| **Uganda** | 17 | 3,413 | 0 |
| **Zambia** | 78 | 2,612 | 0.6 |
| **Zimbabwe** | 32 | 3,561 | 0.3 |

(Source: worldometers)

Although the agriculture and food sectors have been allowed to continue to operate, the World Bank has estimated that the agriculture sector could contract by up to 7%; and that food imports could decline substantially from 13 to 25 percent due to a combination of higher transaction costs and reduced domestic demand [11]. As currencies weaken and the price of food items rise, more and more households will experience hunger and food insecurity.

The impact of rising household poverty and food insecurity will be aggravated further by the psychological and emotional distress accompanying home confinement. Reports of an increase in the incidence of domestic and intimate partner violence points to lockdown measures having a gender dimension [12]. This includes the disproportionate impact of the closure of schools and other educational establishments on girls and women, which will also increase social and health inequalities as children from poorer homes suffer disproportionately from educational deprivation. In some places, the closure of schools will also add to economic and food insecurity amongst the poorest households due to the absence of school meals.

Health may also be harmed as a result of conflict and violence arising from agencies such as the police and army compelling and sanctioning households and communities that are unable or unwilling to comply complying with the measures. In South Africa, both the Human Rights Commission and the Independent Police Investigative Directorate made pointed statements about monitoring and investigating incidents of police misconduct and human rights abuses during the lockdown, [13,14] while in Ghana, the police promised ‘democratic policing strategies’ [15].

Lockdown has also affected the functioning of the health system by increasing physical and financial barriers to accessing healthcare, diverting attention and resources towards COVID-19, and causing patients to stay away from hospitals for fear of contracting COVID-19. A recent study that modelled the impact of COVID-19 on the interruption of HIV/AIDS, TB and malaria services in low and middle-income countries predicted a 10-36% increase in related deaths over a five year period [16]. It was predicted that the greatest impact from HIV would be from interruptions of anti-retroviral therapy; reductions in the timely diagnosis and treatment of new TB cases; and disruptions to insecticide-treated bednet campaigns for malaria. Another concern is the interruption of vaccine delivery due to overstretched healthcare services, parents not bringing their children to clinics because of COVID-19 and disruptions to vaccine supply chains [17].

Lockdown measures can also have negative political consequences that public health agencies should consider. These include the prolonging or disproportionate imposition of restrictions on personal freedoms and civil liberties, as well as the suspension of democratic procedures and safeguards. Several countries have declared states of emergency in response to COVID-19, giving governments extraordinary powers. But such powers should be proportionate to the magnitude and nature of the threat faced by a country, and should ideally be kept under constant review.

Many of the collateral effects of the lockdown are negative. However, there are also positive effects> These include a reduction in green house gas emissions and other pollutants, [18] a lowering in the incidence of traffic accidents, and the creation of more time for some families to be together. In South Africa, the banning of alcohol and tobacco sales have also had positive health impacts. COVID-19 and lockdown also offer a stimulus for governments to reappraise globalised economic, trade and transport systems, animal-based food systems, and ecosystems and biodiversity issues with the aim of improving resilience and reducing risk in relation to the emergence of new infectious disease risks. The interdependence of communities and states, as revealed by COVID-19, also provides an opportunity to reassert social and cultural institutions that encourage trust, respect and tolerance instead of creating fear and suspicion.

## Conclusions

The use of lockdown measures to control COVID-19 is undeniably complex. Not only do the measures have wide-ranging health, social, political and economic effects that produce both benefits and harms, the benefits and harms or lockdown measures are not shared equally across a country’s population. Furthermore, the full impact of lockdown measures will only be known many years into the future. The associated collateral effects of worsening poverty and food insecurity, as well as uncertainty about the feasibility of effective COVID-19 control transmission, raises particular questions about the appropriateness of lockdown in the SSA context. Indeed, in Ghana, there was considerable debate over whether some lockdown measures should have been lifted in the third week of April. This paper highlights the need for inter-sectoral and trans-disciplinary research that is capable of providing a rigorous and holistic assessment of the harms and benefits of lockdown. However, it may be reasonable at this moment in time to suggest that the threats of COVID-19 and the collateral damage of lockdown measures are respectively lower and greater in SSA than in other regions of the world.

## Data Availability

The data presented in this article are publicly available (e.g. Worldometer, World Bank, WHO etc.). For further details please contact the corresponding author

## Acknowledgement

NH, AY, AG, DA, RA, MMAH, DYM,LM, NK, PCK, RK are part of PANDORA-ID-NET Consortium (EDCTP Reg/Grant RIA2016E-1609) funded by the European and Developing Countries Clinical Trials Partnership (EDCTP2) programme which is supported under Horizon 2020, the European Union’s Framework Programme for Research and Innovation.

## Conflict of interest

The authors declare that they have no conflict of interest

## Funding Source

There was no fund for this research.

## Ethical approval

This study does not include any individual level data and thus does not require any ethical approval.

---

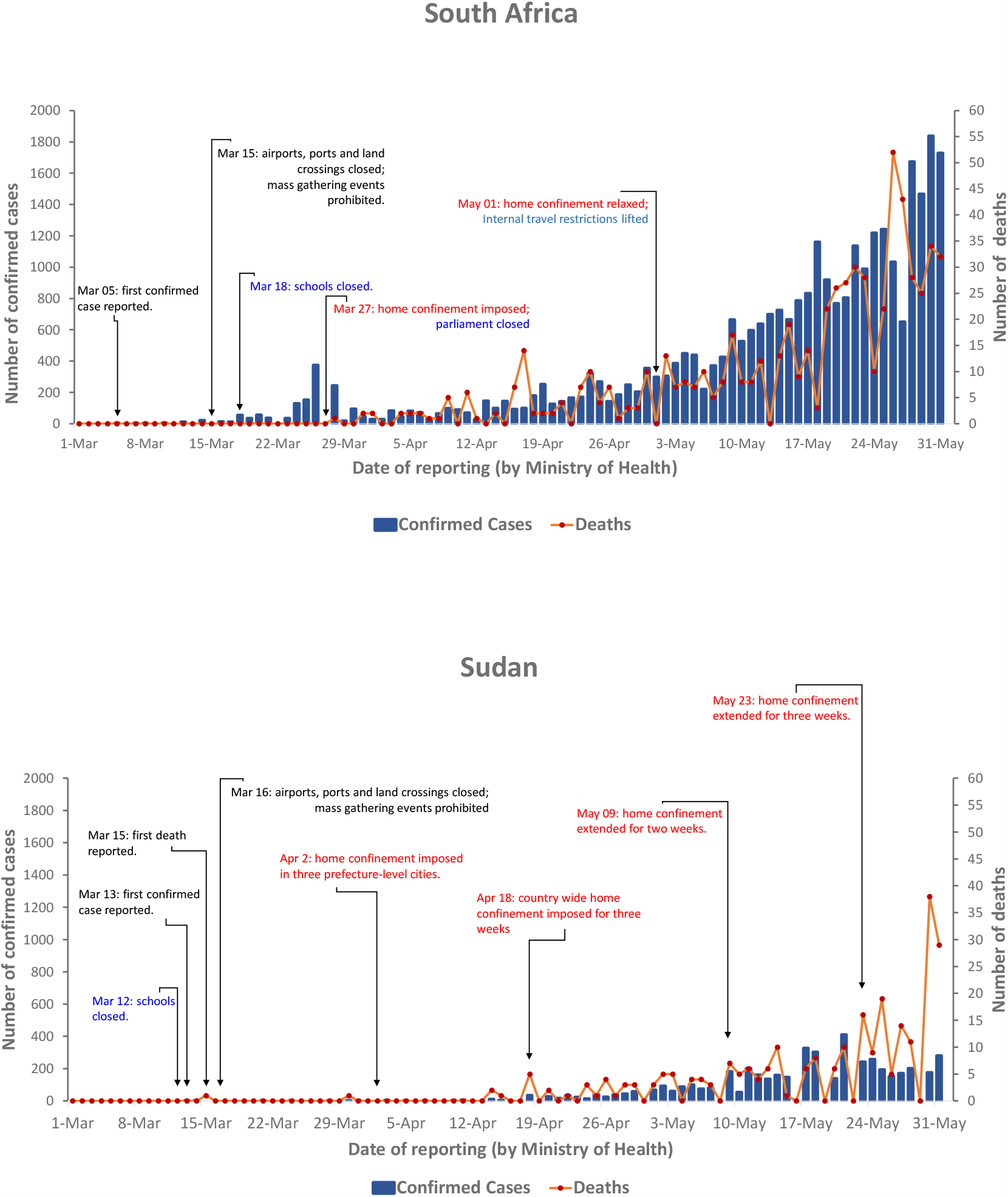

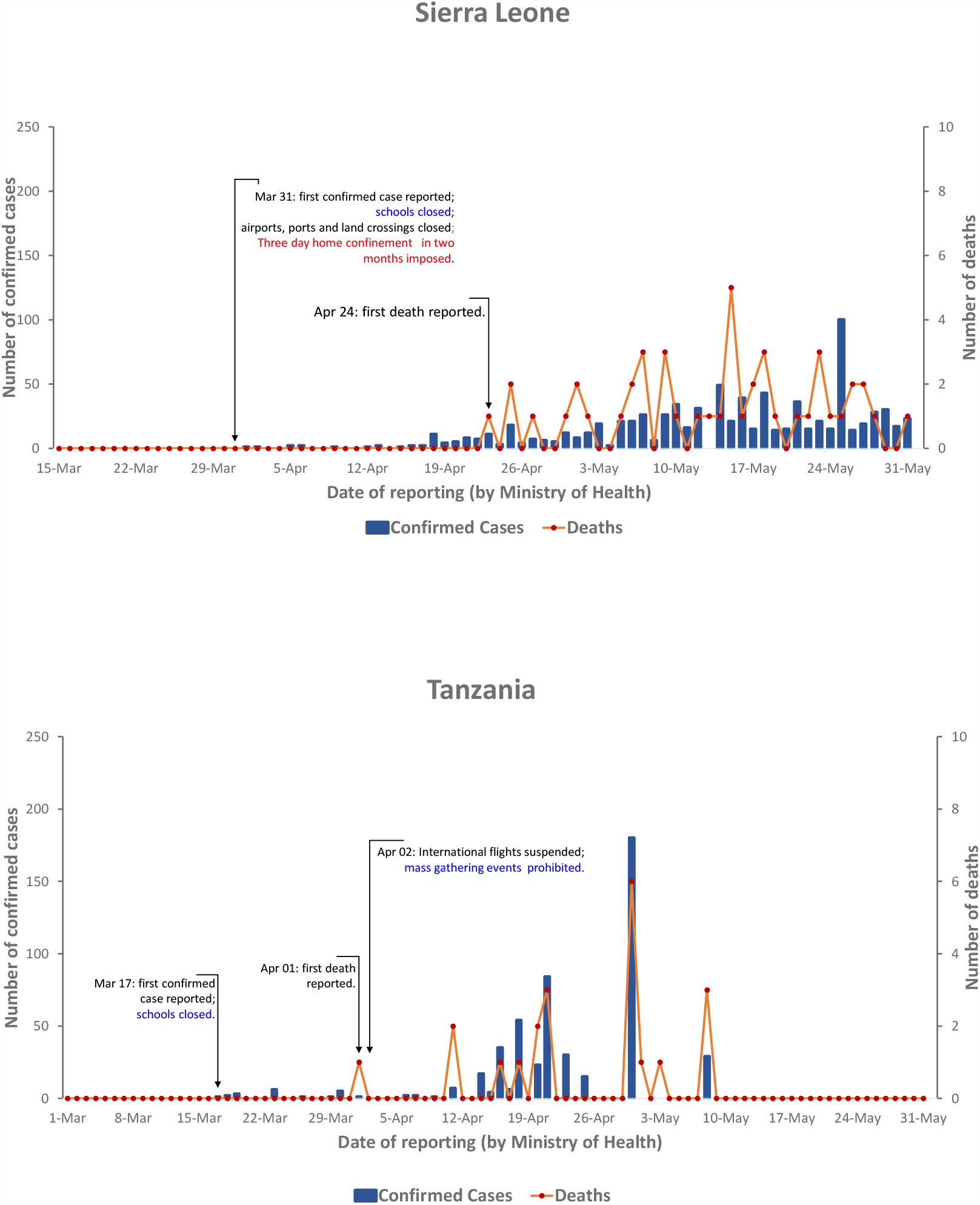

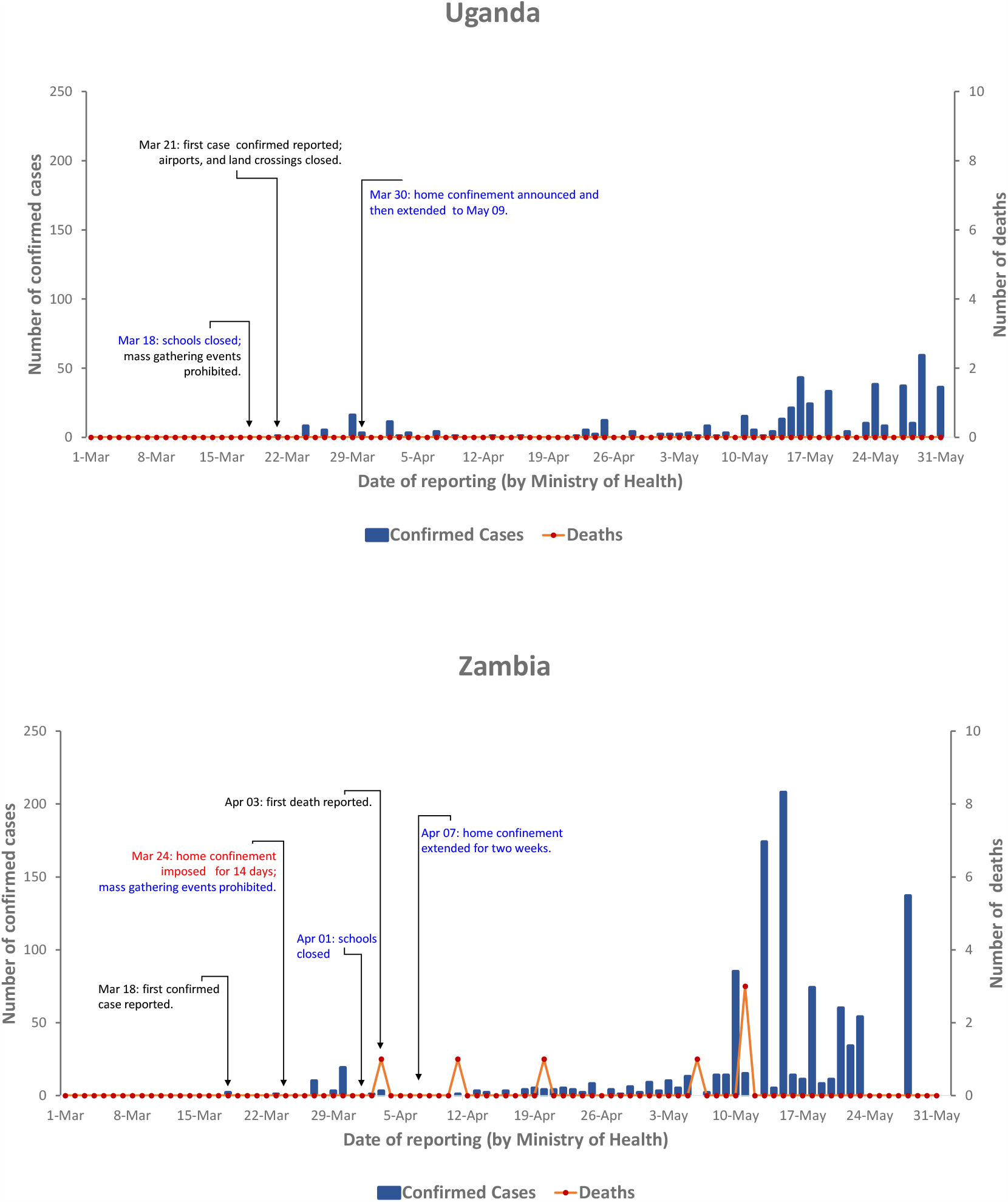

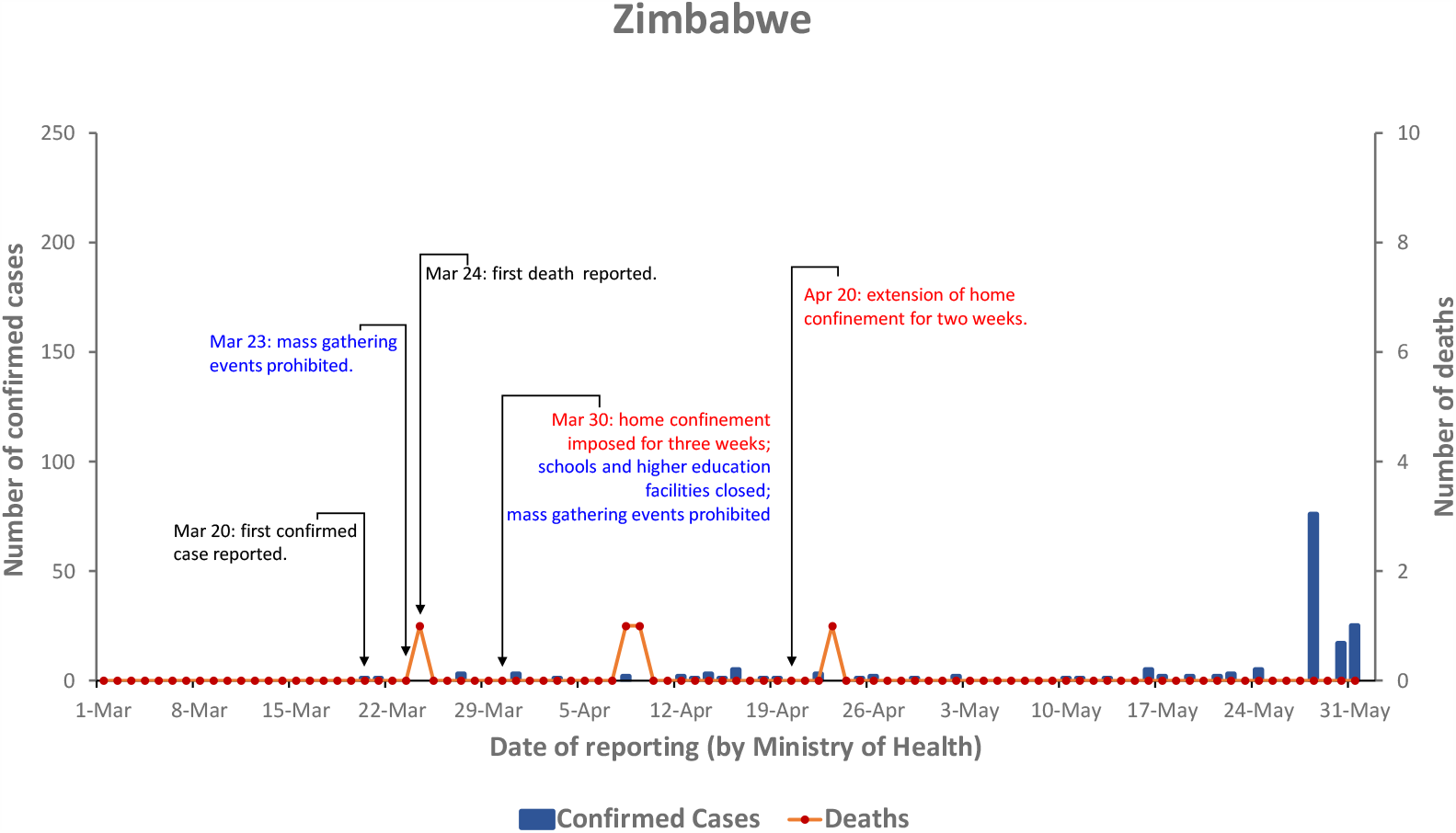

## Notes

### Competing Interest Statement

The authors have declared no competing interest.

### Clinical Trial

NA

### Funding Statement

We received no fund for this research.

### Author Declarations

This study does not contain any personal level data and thus exempted from IRB approval.

